# One Health at the Last Mile: Multi-scale Predictors of *Schistosoma japonicum* Infection in Southwest China across Two Decades of Control

**DOI:** 10.1101/2025.09.23.25335975

**Authors:** William W. Zou, Elise N. Grover, Liu Yang, Elizabeth J. Carlton

## Abstract

In China, schistosomiasis is targeted for elimination. As the country approaches elimination, it is critical to evaluate how the dynamics of transmission are changing in remaining pockets of disease. We have been studying areas of schistosomiasis reemergence and persistence in Sichuan, China since 2007. This study used gradient boosting machines to identify key predictors of infection across two periods, 2007–2010, a period when schistosomiasis had reemerged, and 2016–2019, a period when schistosomiasis was approaching elimination. We also evaluated how key risk factors have shifted over time and whether combinations or predictors amplified risk. We considered predictors describing agriculture, domestic animals, socio-economic status, water and sanitation infrastructure and demographics at individual, household and village-level scales. Our re-emergence and elimination models demonstrated strong predictive performances (AUC-PR=0.92 and AUC=0.85, respectively). In both periods, a person’s age and village level agricultural practices including the average area of dry crops, rice planted, and night soil use, were among the most influential factors. Village-level factors dominated in 2007-2010, while household and individual predictors gained prominence in 2016-2019. Between 2007-2010 and 2016-2019, there were notable increases in the importance of household agricultural practices such as the area of dry crops and rice cultivated, and household cat and dog ownership, while factors describing water and sanitation infrastructure decreased in influence. In the elimination period our models found the combination of high village dry crop cultivation and lack of improved sanitation amplified infection probability. Our findings suggest adding precision interventions targeting high-risk households on top of existing community-wide measures may accelerate schistosomiasis elimination. Practitioners should consider adding agricultural, sanitation and animal infection data to end-game surveillance programs, while researchers validate these patterns in other low-endemic settings and explore causal pathways to inform adaptive, locally tailored strategies.

**Author Summary:** Schistosomiasis is a parasitic disease that has been a target of disease control efforts globally, with China aiming to eliminate the disease. In China, disease control efforts have been successful in reducing the spread and prevalence of the disease, though there are remaining pockets of low levels of transmission. Our study compared the most important factors for schistosomiasis infection risk between two periods, 2007–2010, a period when schistosomiasis had reemerged, and 2016–2019, a period when schistosomiasis was approaching elimination. We found village-level factors were the most important factors behind disease risk in the earlier and later periods, while household and individual-level increased in importance in the later period. Dry crops and rice crop areas at the village-level were also positively associated with disease risk. The importance of potential animal hosts such as ownership of cats and dogs also increased over time. We also found that peak disease risk shifted from 40-60 to >80 years of age. Our results indicate that the factors behind disease may be changing, potentially due to the selective pressures of decades of disease control and largescale socioeconomic changes such as urbanization.

## Introduction

As the World Health Organization (WHO) pushes for global schistosomiasis elimination by 2030, there is an urgency to understand the epidemiological dynamics that characterize the final stages of disease control [1]. The edge of elimination for schistosomiasis remains open and under-explored: do the drivers of transmission shift? Do interventions need to be adapted as prevalence declines? Identifying who remains at risk and which factors sustain transmission is important for effective control efforts in the face of changing ecological, economic, behavioral, and demographic conditions.

China offers a strong case study for investigating these questions. Once hyperendemic for *Schistosomiasis Japonicum*, with a seroprevalence of 34.8% in 1982, China has achieved largescale reductions through a national control program involving mass drug administration, snail control, and targeted treatment of bovine reservoirs [2]. By 2020, seroprevalence had dropped to 2.4%, and the number of endemic counties had declined substantially [3]. Despite this progress, schistosomiasis remains endemic in 354 counties as of 2023, including the rural and mountainous regions of Sichuan province, where seroprevalence remains at 1.24% [4]. While bovines have long been the primary reservoir host targeted for control, emerging evidence suggests that other animals, such as dogs and rodents, may also contribute to ongoing transmission [4–5]. Simultaneously, broader socio-economic shifts, including an aging rural population and land-use change, may be reshaping patterns of exposure in ways that legacy interventions do not fully address [6–7]. Understanding how these factors impact infections, whether their importance has shifted over time, and the extent to which different exposures interact to amplify risk is critical for refining China’s control strategy and for informing global efforts in similar late-stage landscapes.

To capture these evolving dynamics, we used infection, household and demographic data from 2007 to 2019 in rural Sichuan, China and analyzed it using Gradient Boosting Machines (GBMs) to compare predictors of infection during two critical time periods: 2007–2010, when schistosomiasis reemerged in parts of Sichuan [8], and 2016–2019, when the country approached its elimination targets [9]. These methods are well-suited for modeling complex, non-linear relationships and interactions between predictors, building on prior work by Grover et al., and Buchwald et al., [10–12]. Machine learning approaches offer new opportunities to uncover patterns and test how the relevance of environmental, agricultural, WASH, and socio-economic variables has changed in response to decades of control [13]. By evaluating shifts in predictor importance, we provided insight into the changing dynamics of transmission and identified optimal conditions for human infections to occur.

## Materials and Methods

### Study region and design

Data were collected in 2007, 2010, 2016, and 2019 in Sichuan, China, as part of an ongoing study of schistosomiasis reemergence and persistence in areas targeted for elimination. The study has focused on areas where the disease was suspected to be present despite aggressive local control measures, adding new areas of suspected transmission over time. It is not intended to provide a representative sample of Sichuan province. Details of village selection and survey methods have been described in detail elsewhere [11] and in the appendix.

### Human infection data

For each of the study years, infection status was determined by examining up to three stool samples over three consecutive days using the miracidium hatching test, examining 30 grams of stool per sample [8]. In 2007 and 2010 one stool sample was also tested using the Kato-Katz thick smear procedure examining three slides per sample [8]. Infection surveys were conducted in November and December of 2007, 2010 and 2019, and in June and July of 2016 [8]. People were classified as infected if any of the tests were positive. Individuals who tested positive were notified and referred to the local schistosomiasis control station for treatment.

### Ethics statement

This study was approved by the Sichuan Institutional Review Board (China), the University of California, Berkeley (USA), Committee for the Protection of Human Subjects, and the Colorado Multiple Institutional Review Board (USA). All participants provided written, informed consent.

### Infection risk factors

Data on potential risk factors were collected from household and demographic surveys. During the summers of 2007, 2010, 2016 and 2019, the head of each participating household completed a survey with close-ended questions regarding socioeconomic status, domestic and farm animal ownership, sanitation and water access, and agricultural practices. Demographic surveys were conducted as part of the census and administered to the participant directly, or to the head of household. In 2010, demographics surveys were only administered to new participants. All surveys were conducted in the local dialect by trained staff from the Sichuan Center for Disease Control and Prevention and the county Schistosomiasis Control Stations.

We selected candidate predictors of human infection based on six categories that we hypothesized could contribute to human *S. japonicum* infection risk: 1) agricultural practices, 2) animal reservoirs, 3) individual demographics characteristics, 4) occupational risk factors, 5) socioeconomic status (SES), and 6) WASH indicators (Table 1). Additionally, we included a county predictor to account for differences in schistosomiasis control program administration and other differences between counties not captured in the aforementioned risk factors.

**Table 1.**
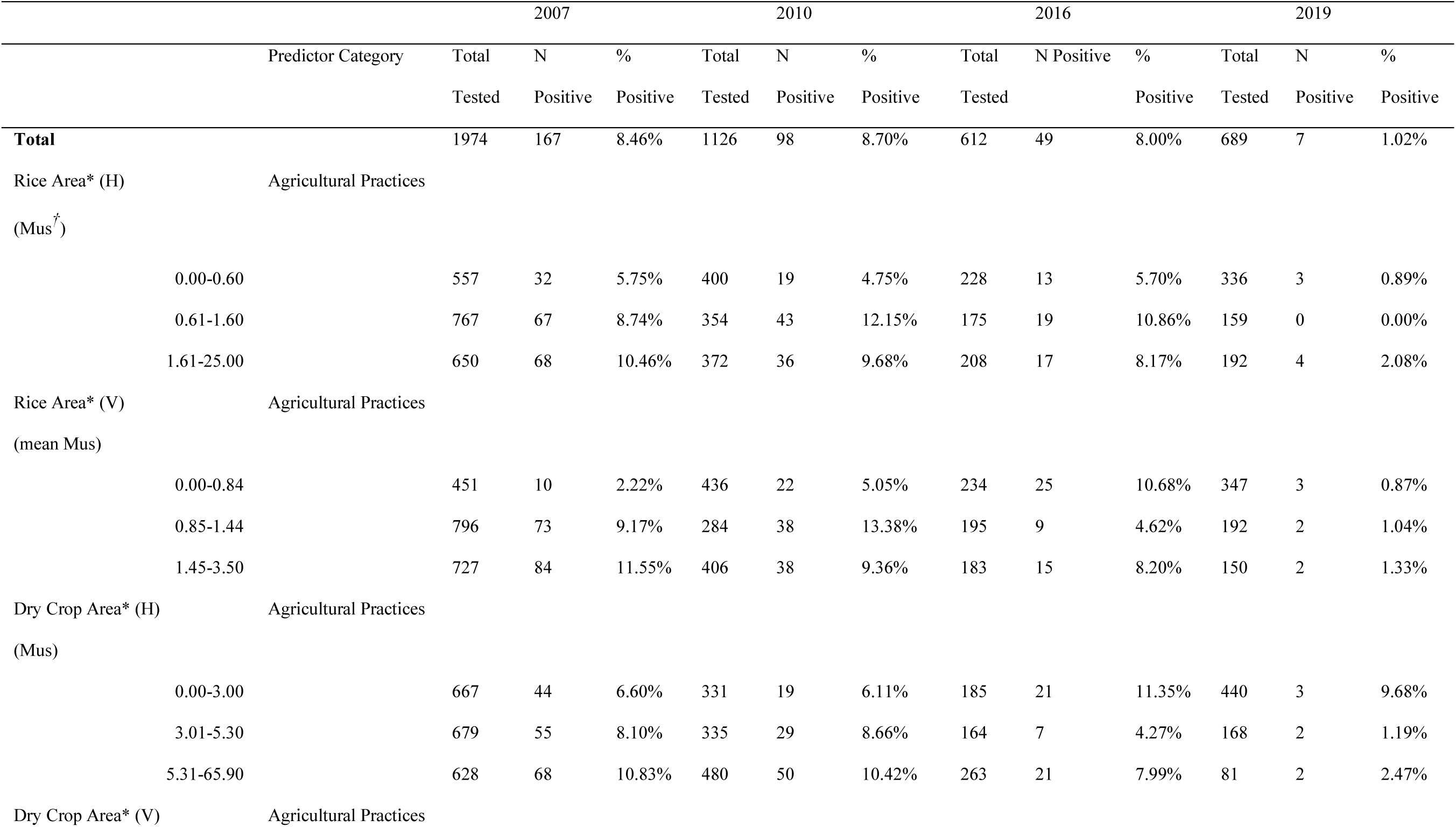
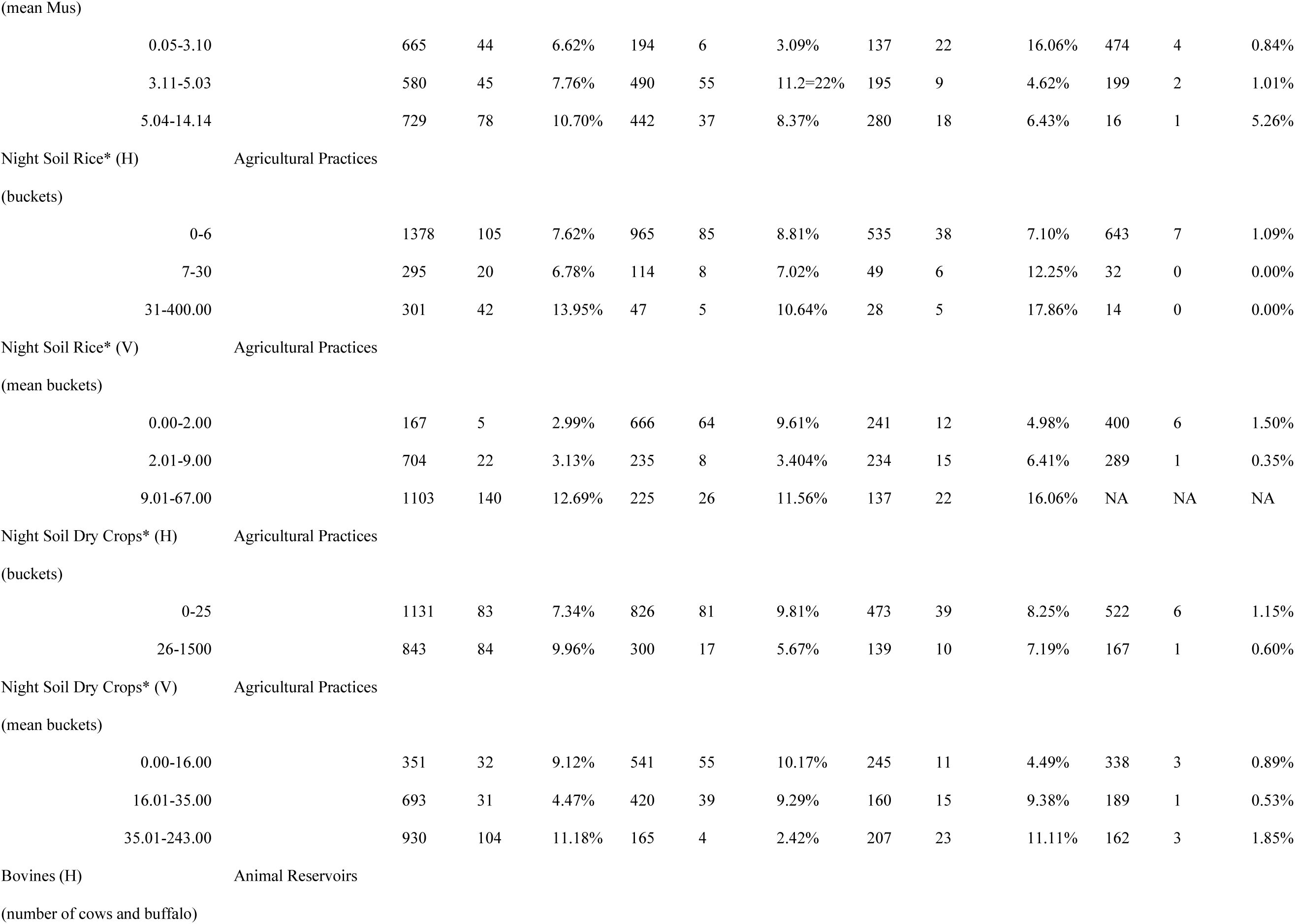
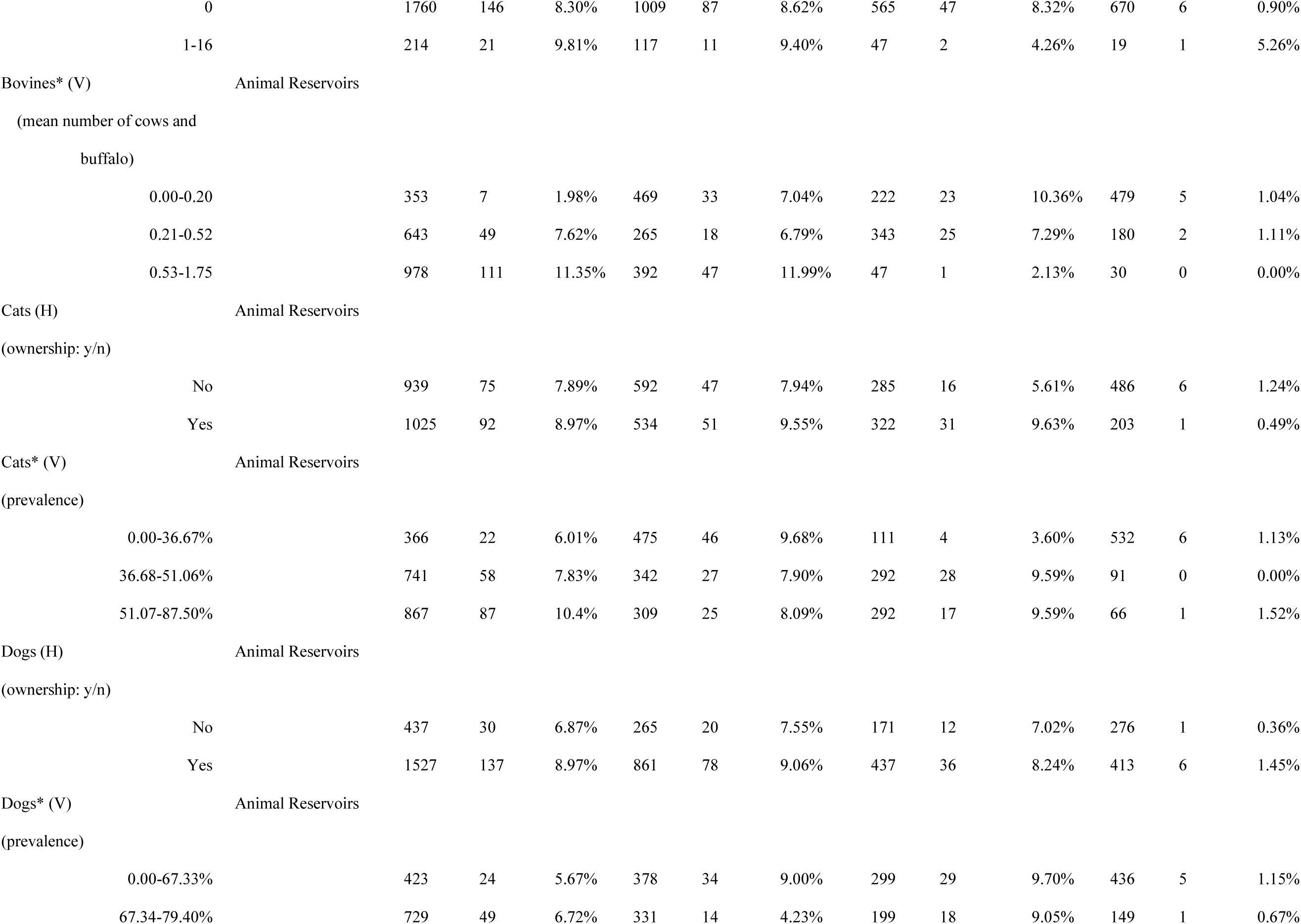
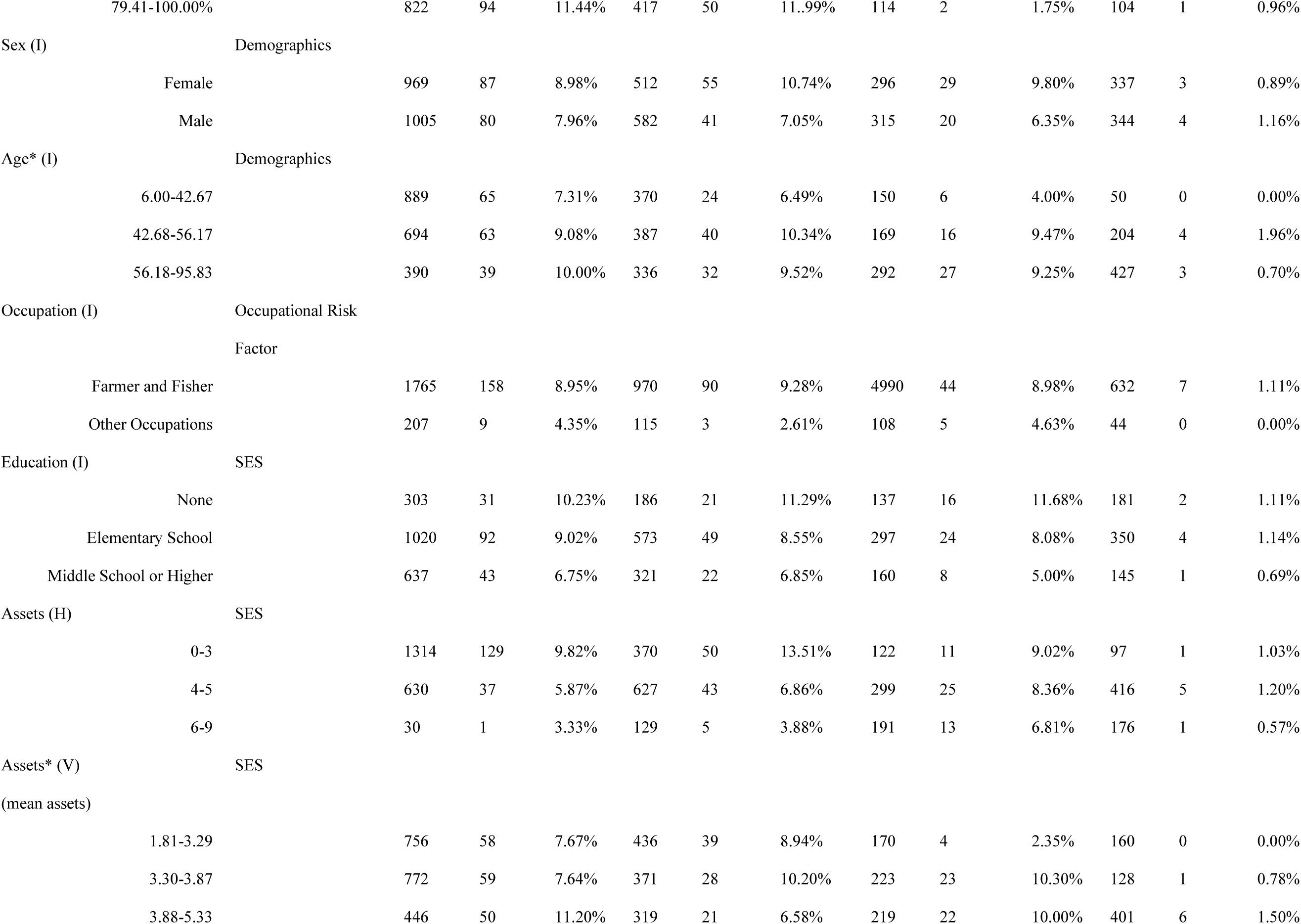
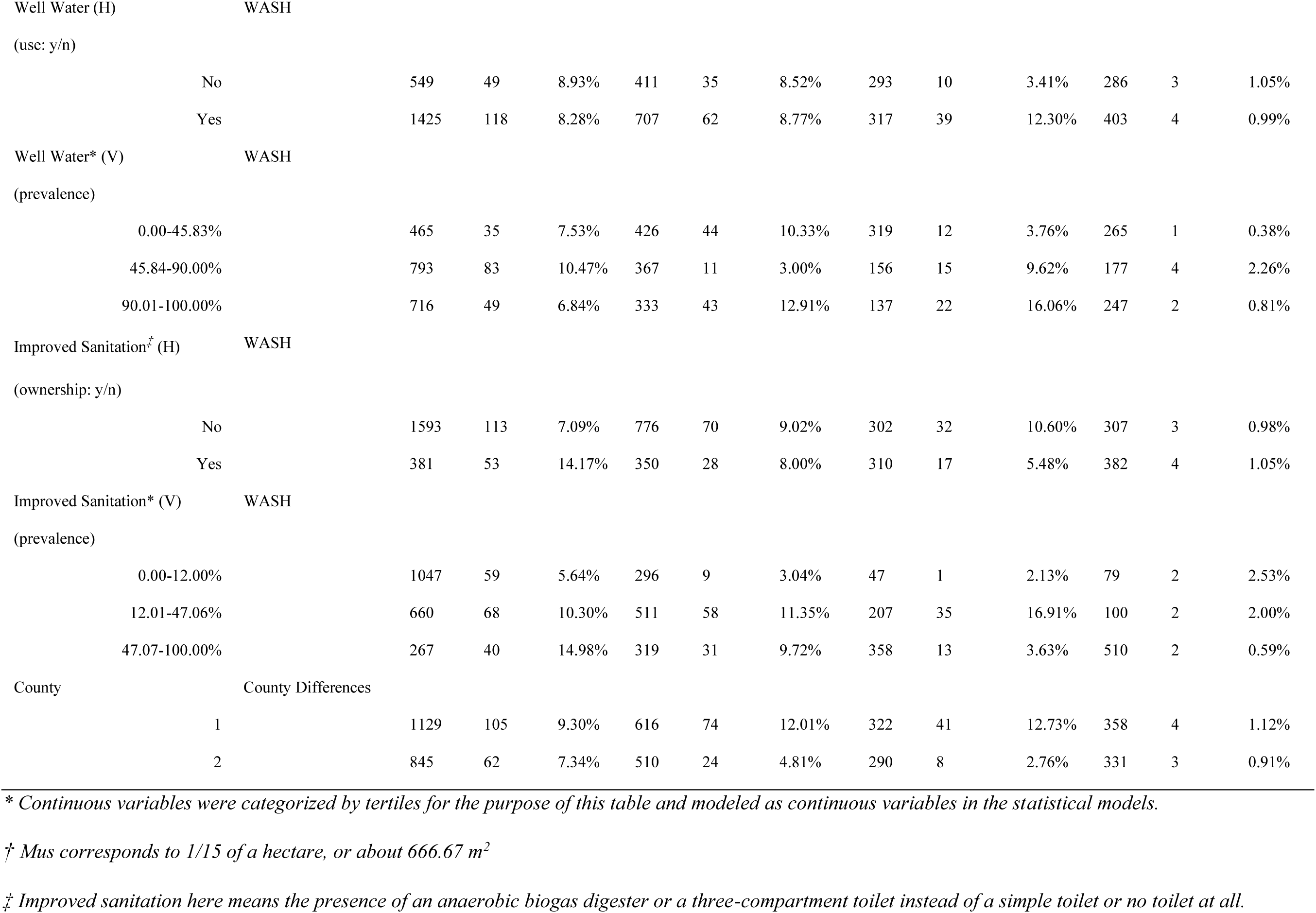
Description of candidate predictors for *S. japonicum* infection by year, including variables that describe agricultural practices, potential animal reservoirs, individual characteristics, occupation, socio-economic status, and access to water, sanitation and hygiene (WASH) infrastructure.

In the study region, common agricultural practices include growing a range of crops throughout the year (e.g., rice, corn, vegetables, wheat, rapeseed) and using night soil (a mix of human and animal waste extracted from stool pits), which is applied as an agricultural fertilizer, in addition to chemical fertilizers. We included estimates of total annual crop area, and the annual volume of night soil applied to crops as predictors of human infection. Because rice farming involves flooding fields in ways that can lead to snail habitat formation and distinct water contact patterns, we also categorized crop area and night soil use into two groups: “rice crops” and “dry crops” (i.e., all other crops).

We generated a nine-point composite asset score as a proxy for SES, following Grover et al. [11]. Household assets were derived from the household survey, wherein the head of household was asked whether they owned any of ‘the following eight assets: tractors, televisions, air conditioners, refrigerators, computers, cars or trucks, motorcycles, and washing machines.

The asset score also included an indicator of whether the home was constructed from either brick or concrete (as opposed to adobe). The composite score was calculated by summing up all reported assets for a given household, yielding a score between zero and nine.

We developed village-level variables that summarized agricultural, animal reservoir, SES and WASH related risks to provide broader context on community-wide exposure and risk factors and capture environmental and socio-economic influences that may impact schistosomiasis infection risk beyond the individual or household scale. Village-level variables were constructed from household survey data collected from all other households located in the same village, excluding data from the household itself to avoid interdependence between household-level and village-level variables. We aggregated continuous household variables to village-mean values and binary household variables to a village-proportion value.

We established criteria for including, removing, or collapsing variables. Only predictors that were available in all study years were included in this analysis. We decided *a priori* to exclude or transform variables with less than 10% or over 90% of observations in a single category. For example, for variables like occupation where the original formulation included several rare (<10%) categories, we opted for a binary formulation of the variable (farmer vs. non-farmer). We also removed highly collinear variables. For example, “House material” was excluded from this analysis because it was already a component of our SES asset score variable. The final dataset included 25 predictors: one county-level, four individual-level, ten household-level, and ten village-level variables (Table 1). Further details of the predictors can be found in table S1 of the appendix.

### Statistical analysis

We used GBMs to identify key predictors of schistosomiasis infection risk, evaluate their relationships with infection outcomes, and assess interactions between predictors. GBMs combine multiple regression trees into an ensemble, enabling them to fit complex, non-linear relationships, which is particularly useful for capturing the non-linear dynamics of disease transmission. Compared to traditional regression models, GBMs can provide stronger predictive performance when modeling biological processes like schistosomiasis transmission, which often involve complex interdependencies [10]. Models were constructed in R using the “gbm” and “dismo” packages [14–15].

We bifurcated the data to evaluate potential changes in infection risk factors between 2007–2010 and 2016–2019. The first period (2007–2010) covers the “reemergence” period, the period shortly after *S. japonicum* reemergence was recognized in the region, and a period when infections were relatively higher (human infection prevalence 8.63% in 2007 and 2010). The second period (2016–2019) covers the “elimination” period, a period when infections were increasingly rare (human infection prevalence 4.51% in 2016 and 2019) and *S. japonicum* elimination was being aggressively pursued.

We partitioned the two datasets into spatially balanced (similar proportion of observations across villages) and temporally balanced (similar proportion of observations across the years) training (70%), evaluation (20%) and test (10%) sets for the analysis to prevent target leakage and improve the generalizability of the results. This was performed using the “BalancedSampling” package in R [16]. We employed 5-fold cross-validation on our training ses so that the model’s performance was assessed across different subsets of the data to reduce the risk of overfitting.

To address class imbalance in our outcome (8.58% infection in the reemergence period; 4.51% infection in the elimination period), we over-sampled the minority class using Random Walk Oversampling (RWO), implemented via the “imbalance” package [17], which generated synthetic infected observations based on the variance and mean of the infected observations.

The datasets also contained missing values across several predictors (3.27% of data were missing due to ambiguous or incomplete responses from survey participants). We addressed missing data in the training datasets using the “randomForest” package [18] to impute missing values based on the median value for continuous variables and the mode value for binary variables.

Optimal model hyperparameters including learning rate, tree complexity, and number of trees were selected using an automated grid search which iterated over one hundred combinations of hyperparameter values and allowed us to identify the strongest combination of hyperparameters for each model. We assessed model performance using Area Under the Precision-Recall (AUC-PR) curves, sensitivity, specificity, accuracy, and kappa values. We evaluated our models based on AUC-PR it is robust to imbalanced datasets [23].

Our primary analytical goal was to rank predictor importance by relative importance, defined as the proportion of the total reduction in squared error that each variable contributes to the models. As secondary goals, we aimed to describe (i) the shape of non-linear marginal effects and (ii) the strength of two-way interactions in relation to *S. japonicum* infection risk.

For the six most influential predictors from each time period, we graphed these functions as Partial Dependence Plots (PDPs) using the pdp package [19]. Uncertainty around each curve was quantified with 95 % confidence bands generated from 1,000 bootstrap replicates of the training data and reflect the range of predicted infection probabilities at each predictor value.

These plots display the average predicted outcome (in this case, probability of infection) as a function of a single predictor, marginalizing the joint distribution of all other variables in the study. The resulting curves reflect nonlinear relationships between our predictors and the marginal probability of infection, as estimated by our BRT models.

We examined pairwise interactions between predictors because transmission dynamics are shaped by complex interdependencies among environmental, socio-economic, and biological factors. Pairwise interactions were quantified using Elith et al.’s customized “Boosted Regression Tree (BRT)” package [10]. All possible pairwise interactions were made available to the model, but they were not all forced into the final fit. Instead, only those splits that lower out-of-bag deviance while applying shrinkage, a small tree depth, and stochastic subsampling were included in the final BRT model. Regularization settings were included to limit model complexity and overfitting and improve generalizability. The most important interactions were identified using interaction size, a value that functions similarly to relative importance in that it quantifies the strength of pairwise interactions based on the additional variance explained by allowing the two variables to interact, beyond their additive effects alone. The interaction sizes are unitless and do not follow a fixed scale, allowing them to identify relatively strong interactions within a single model. However, they are not directly comparable across models or to relative importance scores, as they reflect localized interaction effects rather than global predictive contributions. We selected an interaction size of >5 as a threshold for reporting. We created three-dimensional partial plots using the “BRT” package for the top three most important interactions to examine the direction of these relationships.

## Results

Our analysis included 3,033 observations from people aged 6 to 96 years old across 2 rural counties and 51 villages in Sichuan province, collected from 2007 to 2019. In the study villages, populations ranged from 7-70 households and 12-156 residents. There was a marked decline in infections from 2007 to 2019, even with our sampling strategy focused on areas thought to have ongoing transmission: in 2007, 8.46% of the 1,974 people tested were infected, and by 2019, 1.02% of the 689 people tested were infected (Table 1).

Our models assessing predictors of schistosomiasis infection during the re-emergence and elimination periods demonstrated strong predictive performance, with AUC-PR values of 0.92 and 0.85, respectively. Both models also had high specificity (100%, i.e., 0 false negatives) and accuracy (≥99%) (Fig 1). Model sensitivity and Kappa values were higher for the re-emergence period (Sensitivity = 0.90, Kappa = 0.95) than the elimination period (Sensitivity = 0.85, Kappa = 0.91).

**Figure 1.**
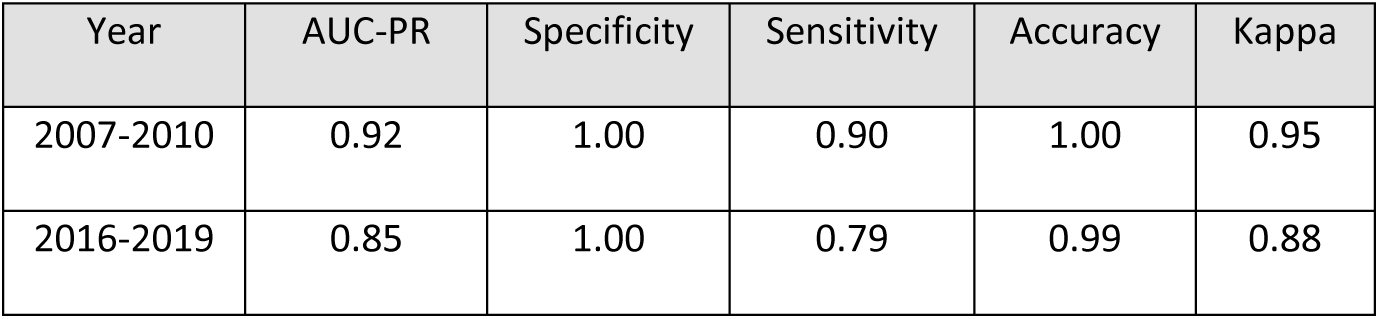
Predictive performance of the reemergence (2007-2010; left) and elimination models (2016-2019; right).

In the reemergence period (2007-2010), the three most important variables (ranked 1-25 with 1 being the most important) were all village-level variables: dry crop area (V), rice area (V) and night soil rice (V) (Fig 2). Age (I), improved sanitation (V), dry crop area (H) and bovine ownership (V) were also strong predictors of infection risk. Of the top seven predictors, five were village-level, one was at the household-level, and one was at the individual-level. The least important were all household-level predictors (improved sanitation, cat ownership, well water usage, and dog ownership).

**Figure 2.**
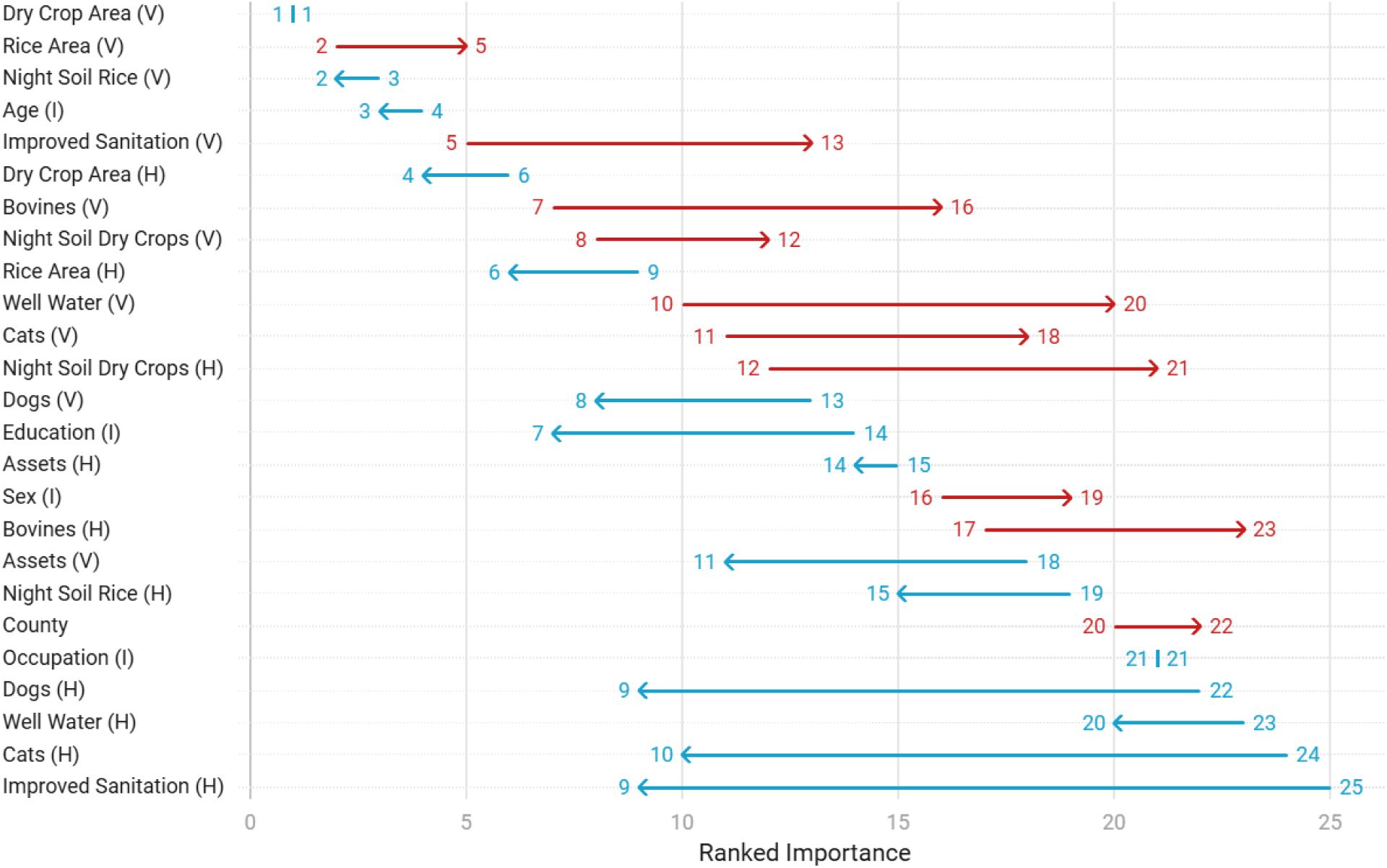
Change in the ranked importance of predictors from 2007-2010 (reemergence period) to 2016-2019 (elimination period).

During the elimination period (2016-2019), dry crop area (V), night soil rice (V), age (I) and dry crop area (H) remained strong predictors of infection (Figure 2). Meanwhile, there were large increases (change of ≥7 ranks) in the ranked importance of cat ownership (H), education (I), and dog ownership (H), assets (V), and improved sanitation (H) all of which were previously ranked low. Of the seven most important predictors, three were village-level, two were household-level, and two were individual-level predictors. The least influential predictors (ranked ≥20) were well water usage (H & V), occupation (I), county, bovine ownership (H), sex (I), night soil dry crops (H), cat ownership (V).

We saw evidence that the relationships between predictors and infection risk displayed a mixture of linear and non-linear effects. In 2007-2010, the marginal change in the probability of infection increased when dry crop area (V) exceeded 8 Mus, or when rice crop area (V) exceeded 2.5 Mus (Fig 3). Infection risk had a positive monotonic relationship with night soil rice (V), rising from an infection probability of 0.01% when 0 buckets of night soil were used on rice crops, to 0.70% when 67 buckets of night soil were used. Infection risk peaked at 0.14% for individuals between the ages of 40-60. Improved sanitation (V) had a net negative, albeit non-linear relationship with infection risk.

**Figure 3.**
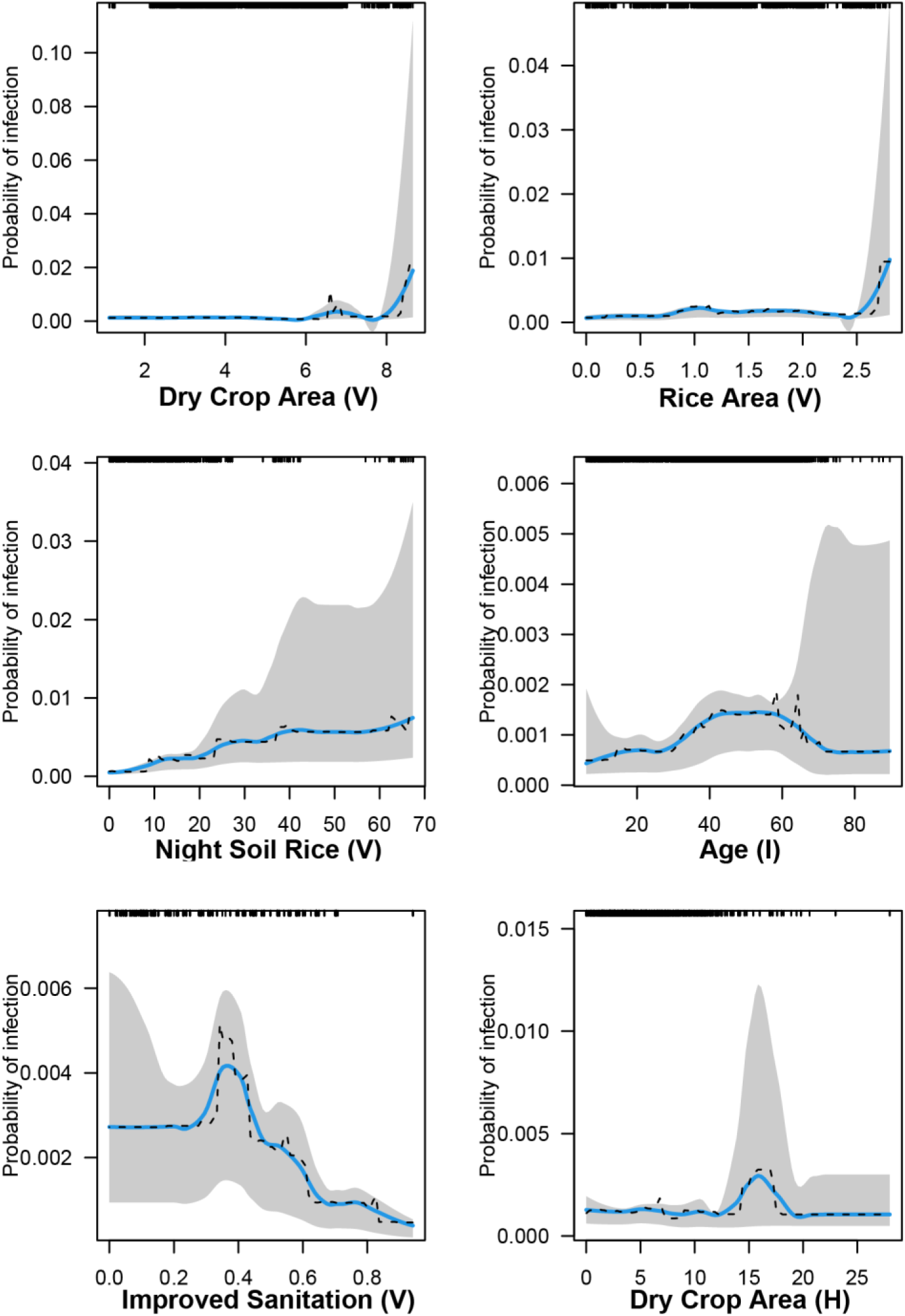
Partial dependence plots (PDP) of the six most important predictors of human schistosomiasis infection risk in 2007-2010 (reemergence model). The PDPs display the change in the average predicted infection risk as predictors vary over their marginal distribution while holding all other variables constant. Fitted curves (dashed lines), smoothing splines (solid blue lines) and 95% confidence intervals based on 1000 bootstrap replicates of the data set (shading) are shown. The full distribution of the predictors is displayed as rug ticks on the top of the plot.

In 2016-2019, infection probability was stagnant when dry crop area (V) and night soil rice (V) were less than 10 and 23 mu respectively, after which point there was a steep increase in infection risk, peaking at 1.7% at 14 Mus of dry crops (V), and 0.43% at 25 mu of rice (Fig 4). Infection risk was also higher in individuals over 80 years of age, and for households planting >10 Mus of dry crops. Larger areas of rice crops at the village and household level also increased infection risk.

**Figure 4.**
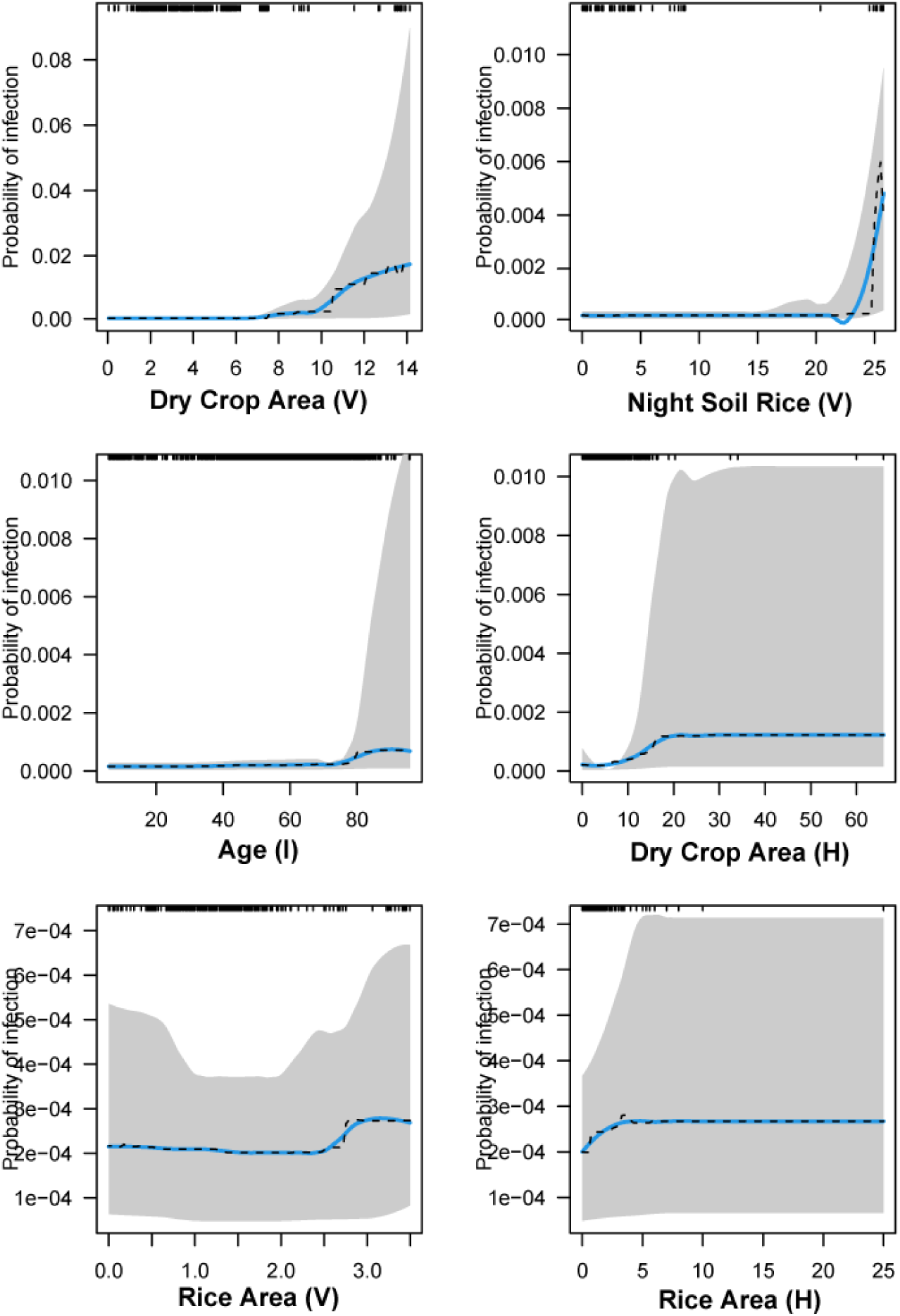
Partial dependence plots (PDP) of the top six predictors of human schistosomiasis infection risk in 2016-2019 (elimination model). The PDP (blue line) displays the change in the average predicted infection risk as predictors vary over their marginal distribution while holding all other variables constant. Fitted curves (dashed lines), smoothing splines (solid blue lines) and 95% confidence intervals based on 1000 bootstrap replicates of the data set (shading) are shown. The full distribution of the predictors is displayed as rug ticks on the top of the plot.

Table 2 presents the strongest pairwise interactions in 2007-2010, showing the combined effects of predictors on human schistosomiasis infection risk measured by their interaction size. The interaction between dry crop area (V) and improved sanitation (V) was the most important interaction, followed by night soil rice crops (V) and dry crop area (H), and Bovines (V) and well water (V). The interaction between dry crop area (V) and improved sanitation (V) had a moderate negative relationship (fig 5). Infection risk was highest for individuals living in households that simultaneously reported planting moderately large areas of dry crops (15 – 17 mu) and were surrounded by households reporting high night soil usage on rice crops (≥60 buckets). Infection risk was also higher for individuals living in villages with a higher presence of bovines (average household ownership >1.5) and higher well water usage (100%).

**Figure 5.**
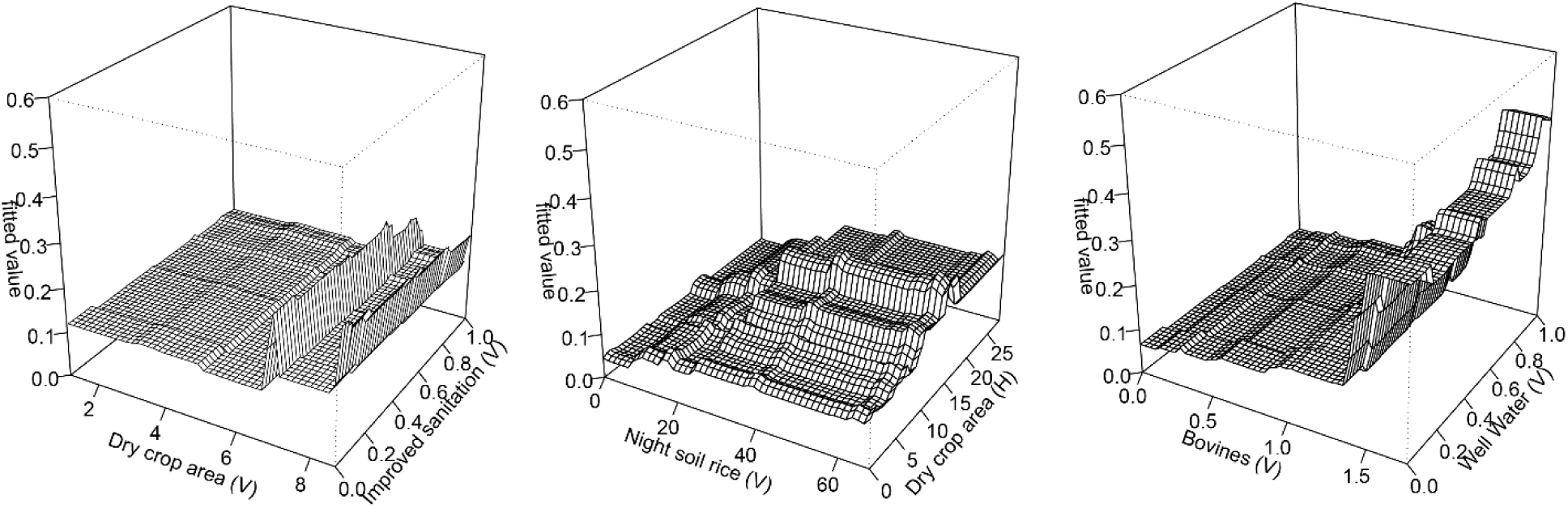
Three-dimensional partial dependence plots of the three most important pairwise interactions in 2007-2010.

**Table 2.**
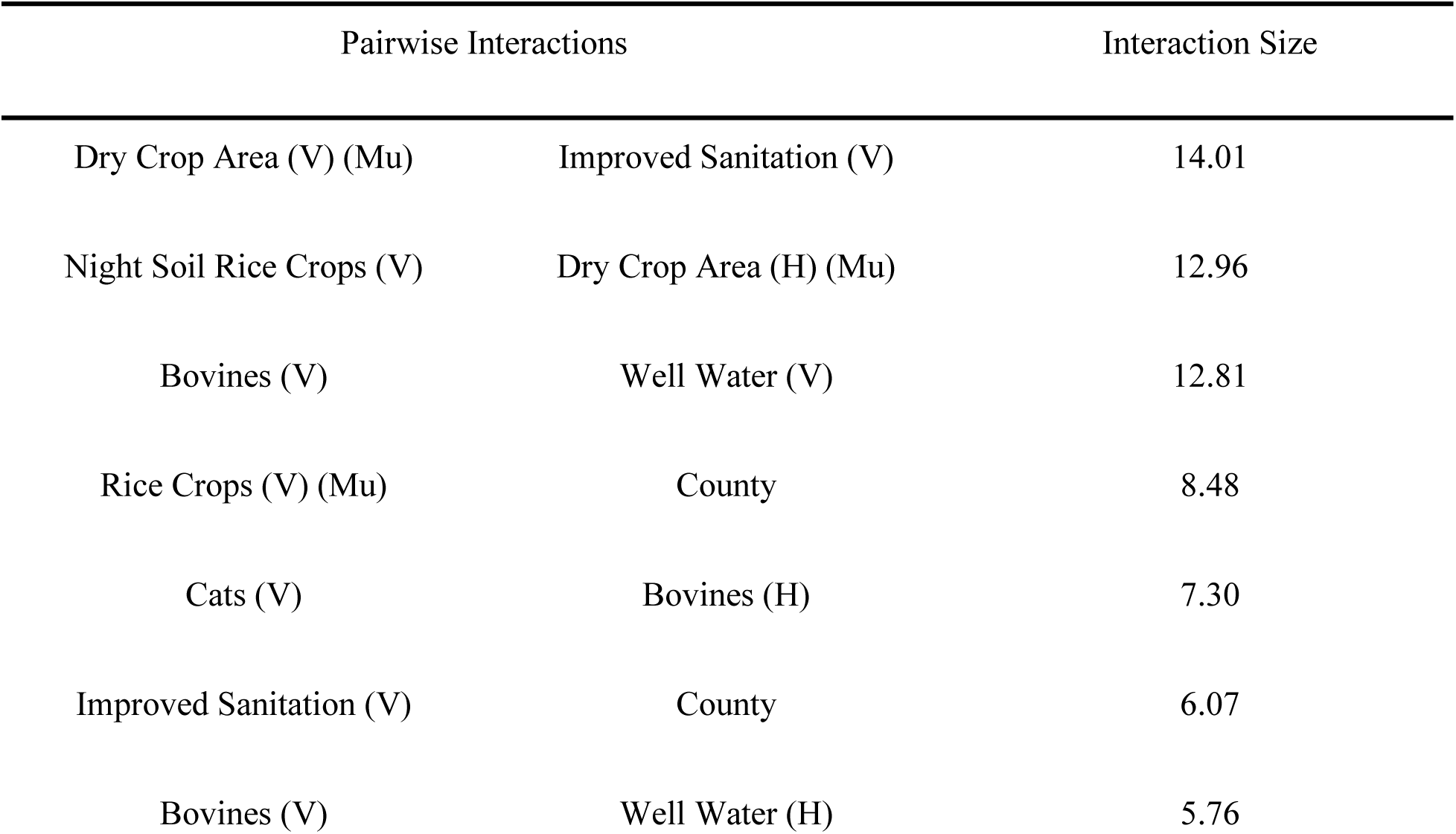
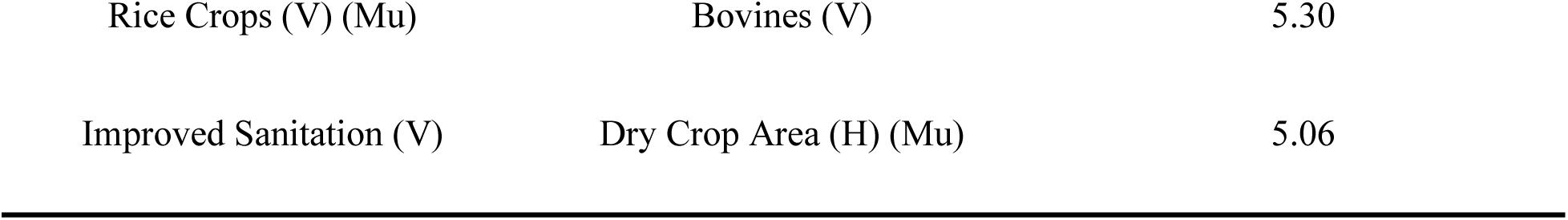
Pairwise interactions with an interaction size > 5 in 2007-2010.

Table 3 presents the highest ranked pairwise interactions in 2016-2019, showing the combined effects of various predictors on human schistosomiasis infection risk, measured by their interaction size. The interaction between the dry crop area (V) and improved sanitation (H) was the most important interaction, followed by dogs (H) and age (I), then dry crop area (V) and age (I). Three of the six most important pairwise interactions included dry crop area (V) and age (I). Infection risk was highest for those over 80 who also owned dogs, or those over 80 who also lived in villages where the reported area of dry crop land was high (>10 mu) (fig 6).

**Figure 6.**
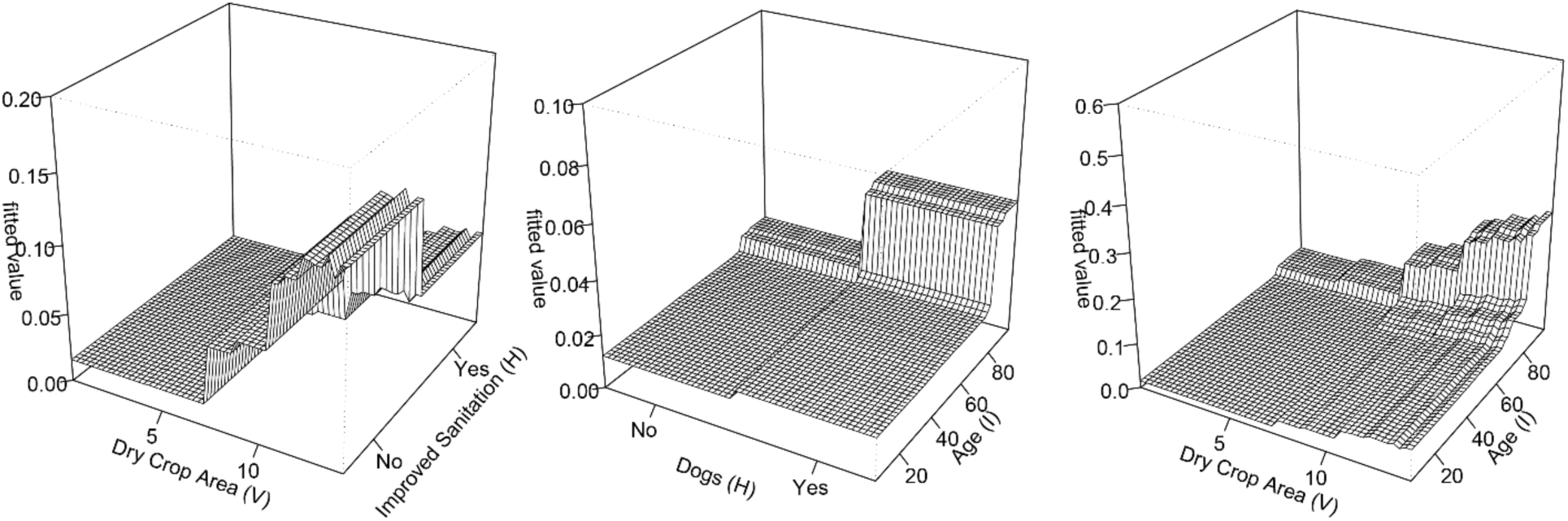
Three-dimensional partial dependence plots of the three most important pairwise interactions in 2016-2019.

**Table 3.** Pairwise interactions with an interaction size > 5 in 2016-2019.

| Pairwise Interactions |  | Interaction Size |
| --- | --- | --- |
| Dry Crop Area (V) (Mu) | Improved Sanitation (H) | 33.36 |
| Dogs (H) | Age (I) | 19.96 |
| Dry Crop Area (V) (Mu) | Age (I) | 18.46 |
| Night Soil Dry Crops (V) | Dogs (H) | 12.54 |
| Night Soil Rice Crops (V) | Dry Crop Area (V) (Mu) | 12.05 |
| Cats (V) | Age (I) | 11.98 |
| Dry Crop Area (H) | Cats (H) | 9.05 |
| Well Water (V) | Cats (H) | 6.27 |

## Discussion

Our findings suggest that the environmental and socioeconomic predictors of *S. japonicum* transmission are dynamic, shifting in importance as the disease moves from reemergence to near elimination, but there was some remarkable consistency in the predictive contribution of agricultural land use. Dry crop area (V), rice area (V), night soil rice (V) consistently emerged as a strong predictor across both time periods, indicating that farming practices maintain a role in disease exposure. Dry crops such as maize, wheat, and rapeseed are often grown on raised embankments or in rotation with paddy rice, creating seasonally moist pockets that may allow snail hosts to persist even when surrounding fields appear dry and may also support other potential reservoirs [20]. Rice paddies, by contrast, requires farmers and livestock to spend prolonged periods in water, increasing exposure opportunities [21]. Night soil may contribute by depositing viable parasite eggs directly into snail habitats while providing soil and water with organic matter that further boosts snail survival [21]. Night soil use emerged as one of the most influential predictors in both the re-emergence and elimination phases of our analysis. Its persistent importance aligns with other studies showing associations between night soil use and human schistosomiasis infections.

While broader indicators of village-level agriculture (e.g., village-level dry crop area) were stronger predictors during the reemergence period, individual and household-level variables (e.g., household-level dry crop area and rice crop area) rose in importance during the elimination period. This trend may reflect a transition from widespread environmental exposure to more specific, household-based transmission risks, suggesting that granular control strategies will be needed as the parasite approaches elimination. The importance of variables describing sanitation also shifted over time. In the reemergence period, village-level improved household sanitation (e.g., the proportion of households in the village with improved toilets) was moderately predictive of infection risk, suggesting that broad community-level improved sanitation access shaped transmission dynamics during this time. However, its importance was markedly lower in the elimination period. In contrast, household-level sanitation (access to a biogas or three-compartment toilet) became a more important predictor during the elimination phase. This shift may reflect a narrowing of transmission pathways: as widespread environmental contamination declines due to village-level improvements, remaining infections may be increasingly influenced by practices and exposure within a given household. It may also suggest that community-level sanitation interventions reach a point of saturation, reducing variation across villages and limiting their predictive value.

The unexpected rise in importance of cat and dog ownership during the elimination period also requires further exploration. One hypothesis is that cats and dogs may play distinct roles in household-level transmission ecology. For example, cat ownership might be protective if they control rodent populations that serve as schistosome reservoir hosts. Recent research indicates that rodents may have become important zoonotic reservoirs of the parasite in endemic regions of China, with increase infection prevalences in mountainous regions such as Sichuan [4]. Conversely, dogs may be associated with higher risk due to their limited role in rodent control and their own susceptibility to *S. japonicum* infection [5]. Dogs have been identified as potential reservoir hosts and may contribute to transmission directly through fecal contamination or indirectly by increasing human contact with water bodies during daily routines such as washing or bathing. Moreover, if households typically own one and not both animals, the presence of dogs may indirectly signal the absence of a potentially protective cat population.

Variables that were most important for predicting schistosomiasis infection at the individual-level were age in the reemergence and elimination period. Age generally had a positive relationship with infection risk in both time periods, but peak risk shifted rightward to older individuals, from a peak risk that occurred at 40-60 years of age to a peak risk that occurred beyond 80 years of age. One possible explanation for this is the rapid urbanization of the country that may have led to younger individuals working in urban areas while older individuals stayed in rural villages to continue farming. The age distribution between the two periods supports this hypothesis, with a more right-skewed distribution in the earlier years and a more left-skewed distribution in the later period (rug ticks, Figures 4-5). Recent literature on the sustainability of farms in China provides further evidence for this finding, with researchers finding that younger farmers were realizing “higher incomes by working in non-agricultural sectors in cities” [22]. The elimination period also saw education become a more important predictor of schistosomiasis infection, with our results suggesting that those who experienced lower to no levels of schooling had a slightly higher infection risk. The mechanism behind this may be that individuals who received less education may be more likely to work in the agricultural sector and thus risk exposure to *S. japonicum* parasites.

Although our models achieved good predictive performance, several limitations merit consideration. First, class imbalance, particularly the small number of infected cases in 2019 (n = 7), could bias estimates and limit generalizability. We addressed imbalance using Random Walk Oversampling, but synthetic examples may not fully capture true heterogeneity in infection risk. Second, while we used cross-validation and bootstrapping to prevent overfitting, the ensemble nature of boosted models means that overfitting cannot be entirely ruled out. Third, missingness in key predictors may have introduced bias despite imputation, particularly in variables derived from self-reported surveys. Fourth, many variables were drawn from household surveys that were administered with changing survey instruments (e.g. different survey questions regarding seasonal crops) over the past decade, although we tried to ensure variable calculations were comparable over the study period. Fifth, infection surveys were collected during the summer of 2016, compared to the winter for all other study years, which may have affected our results since seasonal effects were not accounted for. Finally, the generalizability of our results to other endemic regions is uncertain; Sichuan’s agroecological and control contexts may differ substantially from other settings in China or Southeast Asia. Further validation with external datasets is necessary to assess model generalizability.

Taken together, our findings support the need for adaptive, context-specific control strategies as schistosomiasis moves toward elimination. Agricultural exposures remain central but increasingly heterogeneous, while the importance of sanitation and companion animal presence appears to evolve over time.

## Conclusion

Our findings provide insights into the shifting epidemiology of *S. japonicum* infection in Sichuan province across two different phases of disease control. Our boosted regression models allowed us to explore the interactions between agricultural, socioeconomic, and individual risk factors at multiple spatial scales. The shift from village-level predictors being dominant during the reemergence period to a more balanced distribution of influential predictors across village, household, and individual-levels during the elimination period suggests that as transmission declines, more localized and individualized factors play a greater role in determining infection risk. This aligns with broader trends of urbanization and shifting agricultural practices in China, where younger individuals are leaving rural areas, leading to an older and potentially more exposed farming population.

## Data Availability

The data used in this study contains protected health information (PHI) and cannot be shared publicly due to participant privacy and confidentiality restrictions. Data access for researchers who meet the criteria for access to confidential data may be made available upon reasonable request, subject to institutional and ethical approvals. Requests for data access should be directed to the Colorado Mulitple Institutional Review Board (COMIRB)'s Director, John Heldens.

