## Appendix for "One Health at the Last Mile: Multi-scale Predictors of *Schistosoma japonicum* Infection in Southwest China across Two Decades of Control"

**S1 Appendix/Supporting Information**

**Sampling Strategy**

In 2007 we selected 53 villages from three counties in Sichuan where schistosomiasis had reemerged. A magnitude 7.9 earthquake in 2008 impacted one of the three study counties, leading us to discontinue research in this county. Observations from this county were not included in any of our models. In 2010, the 36 villages in the remaining two counties were surveyed. In 2016, surveillance records were reexamined to focus surveys on the highest-risk locations within the two counties, as schistosomiasis infections were declining. A total of 10 villages were selected for the 2016 survey, including 7 from the 2007 and 2010 surveys, and 3 that were newly added. In 2019, we surveyed 25 villages in the two counties including 9 of the villages surveyed in 2016, and 16 villages where surveillance records indicated possible ongoing or recent schistosomiasis transmission (4 of the these were surveyed in 2007 and 2010). In each village and timepoint, we conducted a panel survey, including a census of all residents aged 6 and older, collecting demographic data, and recruiting these individuals for *S. japonicum* infection testing, and conducting household surveys.

**Table S1. Summary of Predictors Included in the Analyses**

**Table S1.** Summary of predictors included in the analysis

| Predictor | Detailed description (units) | Scale | Category | Variable Type | Notes |
| --- | --- | --- | --- | --- | --- |
| Rice area (H) | Area of rice cultivated by the household in the past year (mu) | Household | Agriculture | Continuous | Almost all rice is cultivated in the summer season. |
| Rice area (V) | Mean area of rice cultivated per household (mu) | Village | Agriculture | Continuous |  |
| Dry crop area (H) | Total area of dry crops cultivated by the household in the past year (mu) | Household | Agriculture | Continuous | Most common dry crops are corn in the summer months, wheat and rapeseed in the winter months. Various vegetables are grown in both winter and summer seasons. |
| Dry crop area (V) | Mean area of dry crops cultivated per household (mu) | Village | Agriculture | Continuous |  |
| Night soil rice (H) | Quantity of night soil applied to rice crops (buckets) | Household | Agriculture | Continuous |  |
| Night soil rice (V) | Mean volume of night soil applied to rice crops per household (buckets) | Village | Agriculture | Continuous | This represents the frequency of night soil x the buckets of night soil used for each instance. |
| Night soil dry crops (H) | Quantity of night soil applied to dry crops (buckets) | Household | Agriculture | Continuous | See explanation for *Night Soil Rice (H)* |
| Night soil dry crops (V) | Mean volume of night soil applied to dry crops per household (buckets) | Village | Agriculture | Continuous |  |
| Bovines (H) | Number of bovines owned by household | Household | Animal Reservoir | Continuous | Surveys allowed respondents to input any integer value of water buffalo and cows they owned. |
| Bovines (V) | Percent of households that own bovines | Village | Animal Reservoir | Continuous |  |
| Cats (H) | Household owns cats | Household | Animal Reservoir | Binary |  |
| Cats (V) | Percent of households that own cats | Village | Animal Reservoir | Continuous |  |
| Dogs (H) | Household owns dogs | Household | Animal Reservoir | Binary |  |
| Dogs (V) | Percent of households that own dogs | Village | Animal Reservoir | Continuous |  |
| Sex (I) | Participant sex | Individual | Individual risk factor | Binary | 0 = female; 1 = male |
| Age (I) | Participant age | Individual | Individual risk factor | Continuous |  |
| Education (I) | Highest level of schooling completed | Individual | Individual risk factor | Ordinal | 1. None  2. Elementary School  3. Middle School or Higher |
| High-Risk Occupation (I) | High-risk occupation | Individual | Occupational risk factor | Categorical | 0. Not farmer  1. Farmer or Fisher |
| Assets (H) | Number of household assets owned | Household | SES | Discrete | One point each is assigned for ownership of nine items including computers, televisions and trucks. One point for living in a brick or concrete home (vs. adobe) |
| Assets (V) | Mean number of assets owned per household | Village | SES | Continuous |  |
| Well water (H) | Household uses well water | Household | WASH | Binary | Minor variations in questionnaire over time^*^ |
| Well water (V) | Percent of households that use well water | Village | WASH | Continuous |  |
| Improved sanitation (H) | Household has a biogas and/or 3-compartment toilet | Household | WASH | Binary | Minor variations in questionnaire over time^†^ |
| Improved sanitation (V) | Percent of households with improved sanitation | Village | WASH | Continuous |  |
| County (C) | County of residence | County | County | Binary |  |

SES: Socio-economic Status

WASH: Water, sanitation and hygiene

* In 2007, the survey directly asked if a household had a well (“well water”), but missing responses were relatively high (N=80). The 2007 survey also included questions as to whether people used well water for household activities such as cooking, washing, or bathing. These responses were used to create a derived "welluse" variable. Missing 2007 “well water” values were filled using the "welluse" variable, making 2007 data comparable with later years. From 2010 to 2019, the question was changed to “Does your household get water from a well?”, which had fewer missing responses and was used to classify household well water use.
